# The role of age in choosing high-efficacy treatment for multiple sclerosis – an Austrian MS Database study

**DOI:** 10.1101/2025.07.14.25331511

**Authors:** Harald Hegen, Fabian Föttinger, Janette Walde, Klaus Berek, Maria Martinez-Serrat, Anna Damulina, Nik Krajnc, Markus Ponleitner, Franziska Di Pauli, Christian Enzinger, Florian Deisenhammer, Thomas Berger, Michael Khalil, Gabriel Bsteh, the Austrian Multiple Sclerosis Database Study group

**Author notes:** Corresponding author: Prof. Gabriel Bsteh, PD, MD, PhD, MSc, Department of Neurology, Medical University of Vienna, Vienna, Austria Waehringer Guertel 18-20, 1090 Vienna, Austria.

## Abstract

**Background:** Treatment strategy for relapsing multiple sclerosis (RMS) is increasingly shifting towards first-line use of high-efficacy DMT (H-DMT). However, DMT efficacy declines with increasing age and the benefit of first line H-DMT at higher age remains unclear. Here, we aimed to investigate whether the superiority of H-DMT over moderate-efficacy DMT (M-DMT) depends on age.

**Methods:** Using the Austrian MS database, we included previously DMT-naïve RMS patients aged ≥18 years, who i) initiated a DMT continuing it for ≥12 months, ii) had MRI at baseline, and iii) had clinical follow-up for ≥24 months. Cox regression analyses including age and DMT strategy (H-DMT vs. M-DMT) plus an interaction effect were employed to predict time to relapse.

**Results:** A total of 215 RMS patients (median age of 41 years [25^th^-75^th^ percentiles: 32-53], 66% females) were observed over a median of 42 (28-58) months. During this period, eighty-one (38%) patients had a relapse. While increasing age was associated with decreased risk of relapse (hazard ratio (HR) 0.95 per year, 95% confidence interval [CI]: 0.93-0.98, p<0.001), the use of H-DMT lowered the risk of relapse compared to M-DMT (HR 0.06, 95%-CI: 0.01-0.45, p=0.007). In patients with H-DMT, the benefit of treatment was reduced by increasing age (HR: 1.06, 95%-CI: 1.01-1.11, per year, p=0.031). Superiority of H-DMT over M-DMT was estimated to be lost at the age of approximately 50 years.

**Conclusion:** The benefit of H-DMT over M-DMT as first-line treatment decreases with increasing age and seems to vanish in patients above approximately 50 years.

**What is already known on this topic:** High-efficacy disease-modifying treatments (H-DMTs) are increasingly used as first-line therapy in relapsing multiple sclerosis (RMS) due to their superior effectiveness in reducing inflammatory disease activity. However, both clinical and radiological disease activity naturally decline with age, and prior meta-analyses suggest that the relative benefit of H-DMT over moderate-efficacy DMTs (M-DMTs) diminishes in older patients. These findings have largely been derived from clinical trials with restricted age ranges and enriched disease activity, limiting their generalizability to real-world, treatment-naïve populations across the full adult age spectrum.

**What this study adds:** In a real-world, national cohort of DMT-naïve RMS patients across a wide range of age, this study shows that while H-DMTs significantly reduce the risk of relapse compared to M-DMTs, their superiority is progressively attenuated with advancing age. Notably, the benefit of initiating H-DMTs as first-line therapy becomes statistically indistinguishable from M-DMTs around the age of 50 years. These findings were independent of baseline disease duration and other covariates, emphasizing age as a key modifier of treatment effect.

**How this study might affect research, practice or policy:** These findings support the integration of age as a critical factor in guiding first-line DMT decisions for RMS. For patients over 50 years, M-DMTs may offer a more appropriate initial treatment option, minimizing exposure to the higher risk profiles of H-DMTs in the absence of clearly superior efficacy. This study underscores the importance of personalized treatment approaches and highlights the need for future clinical trials to include broader age ranges, facilitating evidence-based, age-adjusted treatment strategies in multiple sclerosis.

## Introduction

Multiple sclerosis (MS) is a chronic autoinflammatory demyelinating disorder of the central nervous system (CNS) carrying a high risk of permanent disability ^1^. In relapsing MS (RMS), the advent of disease-modifying treatments (DMT), particularly those with high-efficacy (H-DMT), has enabled a focus on sustained suppression of inflammatory disease activity. Thus, treatment paradigms are increasingly shifting toward the early use of H-DMT, often even as a first-line option, rather than starting with moderate-efficacy DMT (M-DMT) ^2^.

However, it is well established that clinical and radiological inflammatory disease activity, i.e. relapses and new magnetic resonance imaging (MRI) lesions, peak in younger patients and gradually decline with advancing age ^3–6^. In parallel, the comparative efficacy of DMT is most pronounced in younger patients and appears to decline with increasing age in randomized controlled trials, where a meta-analysis reported that the relative benefit of H-DMT over M-DMT vanishes in patients over the age of 40-50 years ^7–9^. Nevertheless, clinical trials selected patients within a narrow age range and enriched for baseline disease activity regardless of disease duration and previous DMT, thereby not fully representing real-world RMS patients ^10^. Hence, age-dependent differences in DMT efficacy are not well-established and the benefit of using H-DMT as a first-line option in RMS patients at higher age currently remains unclear.

In this study, we aimed to investigate in a real-world cohort of DMT-naïve patients, whether the superiority of H-DMT over M-DMT in reducing the risk of relapse depends on age, and, if so, whether there is an age at which the superiority becomes statistically insignificant.

## Methods

### Study design and patients

This is a retrospective cohort study within the Austrian MS database (AMSD) ^11^. Between May and September 2024, the AMSD was screened for patients diagnosed with RMS according to current McDonald diagnostic criteria aged ≥18 years, who i) were newly (i.e. previously DMT-naïve) initiated on a DMT continuing it for ≥12 months, ii) had MRI at baseline, and iii) had a clinical follow-up for ≥24 months (in patients without relapses, or ≥12 months in patients with relapses occurring at least 6 months after DMT start) ^12–14^.

### Research question

Is the superiority of first-line H-DMT over M-DMT in reducing the risk of future clinical disease activity dependent of age?

### Endpoints

The primary endpoint was the occurrence of a relapse after DMT initiation. The secondary endpoint was disability accrual after DMT initiation.

### Definitions

A relapse was defined as patient-reported symptoms and objectively confirmed neurological signs typical of an acute CNS inflammatory demyelinating event with duration of at least 24 hours in the absence of fever or infection and separated from the last relapse by at least 30 days ^14^. Disability accrual was defined as an increase of Expanded Disability Status Scale (EDSS) score from baseline of at least 1.5 points if baseline EDSS was 0, 1.0 point if baseline EDSS was 1.0–5.0, and 0.5 points if baseline EDSS was ≥5.5 confirmed after 6 months ^15^.

The number of hyperintense lesions on T2-weighted MRI (T2L) and the presence of contrast-enhancing lesions on T1-weighted MRI (CEL) were retrieved from the AMSD. MRI scans were done on 1.5 or 3T MR scanners and rated by local experienced neuroradiologists. MRI protocols included 3D fluid attenuated inversion recovery sequences (FLAIR), T2 sequences and T1 sequences.

M-DMT comprised interferon-beta, glatiramer acetate, teriflunomide and dimethyl fumarate. H-DMT included the sphingosine-1-phosphate receptor modulators (fingolimod, ozanimod), cladribine, natalizumab, alemtuzumab and anti-CD20 monoclonal antibodies (rituximab, ocrelizumab, ofatumumab).

### Statistical analysis

Statistical analysis was performed using R ^16^. Categorical variables are shown as frequencies and percentages, continuous variables are displayed either as mean and standard deviation (SD), or median and 25^th^-75^th^ percentiles, as appropriate. Univariate comparisons were done by χ^2^ test, Fisher’s exact test and Mann-Whitney U test.

Multivariable Cox regression was employed using time to relapse (or observation time in stable patients) as dependent variable as well as DMT strategy (binary: M-DMT vs. H-DMT) and age (continuous) as independent covariates (plus an interaction effect) adjusting for sex (binary), disease duration (continuous), EDSS score at baseline (continuous), number of relapses in the prior year (continuous), number of T2L at baseline (binary: <9/ ≥9) as well as presence of CEL at baseline (binary: 0/ ≥1). Similarly, Cox regression was deployed with time to disability accrual (or observation time in stable patients).

To visualize the effect of age and DMT, we computed the estimated Cox regression survival probabilities separately for M-DMT and H-DMT and for different age groups (20, 30, 40 and 50 years, respectively) with sex set to “female”, T2L to “≥9”, CEL to “≥1” and all other parameters to their median values. A p-value <0.05 was considered statistically significant.

An a priori power analysis for the Cox regression regarding the primary endpoint with a significance level of 5%, a power of 80%, and assumptions of i) a hazard ratio of 0.6, ii) a proportion of patients with relapse of 0.4, and iii) a shorter observation time for patients with relapse (ratio of observation time of patients with and without relapse of 0.66) indicated a necessary sample size of 200 patients.

### Ethics

The study was approved by the ethics committee of the Medical University Vienna (ethical approval number: 1668/2023). As datasets were obtained in routine practice and exported pseudonymously, the need for written informed consent from study participants was waived by the ethics committee. This study adheres to the reporting guidelines outlined within the ‘Strengthening the Reporting of Observational Studies in Epidemiology’ (STROBE) Statement.

## Results

A total of 215 patients with RMS at a median age of 41 (25^th^-75^th^ percentiles: 32-53, range: 21-73) years and a female predominance (66%) were included into the study. The detailed inclusion/exclusion process is given in Figure 1. Patients had a median disease duration of 2 (25^th^-75^th^ percentiles: 1-7) months, showed a median EDSS score of 1.0 (0-2.0) at baseline, and had 1.1 ± 0.5 relapses in the 12 months prior to DMT initiation. The T2L at baseline was ≥9 in 58% of patients, CEL were present in 33% of patients. Seventy-four percent of patients started M-DMT, while 26% started H-DMT. The median year of DMT start was 2018 (2016-2020). Detailed demographic, clinical and imaging characteristics of the cohort are given in Table 1 (and eTables 1 and 2).

**Figure 1.**
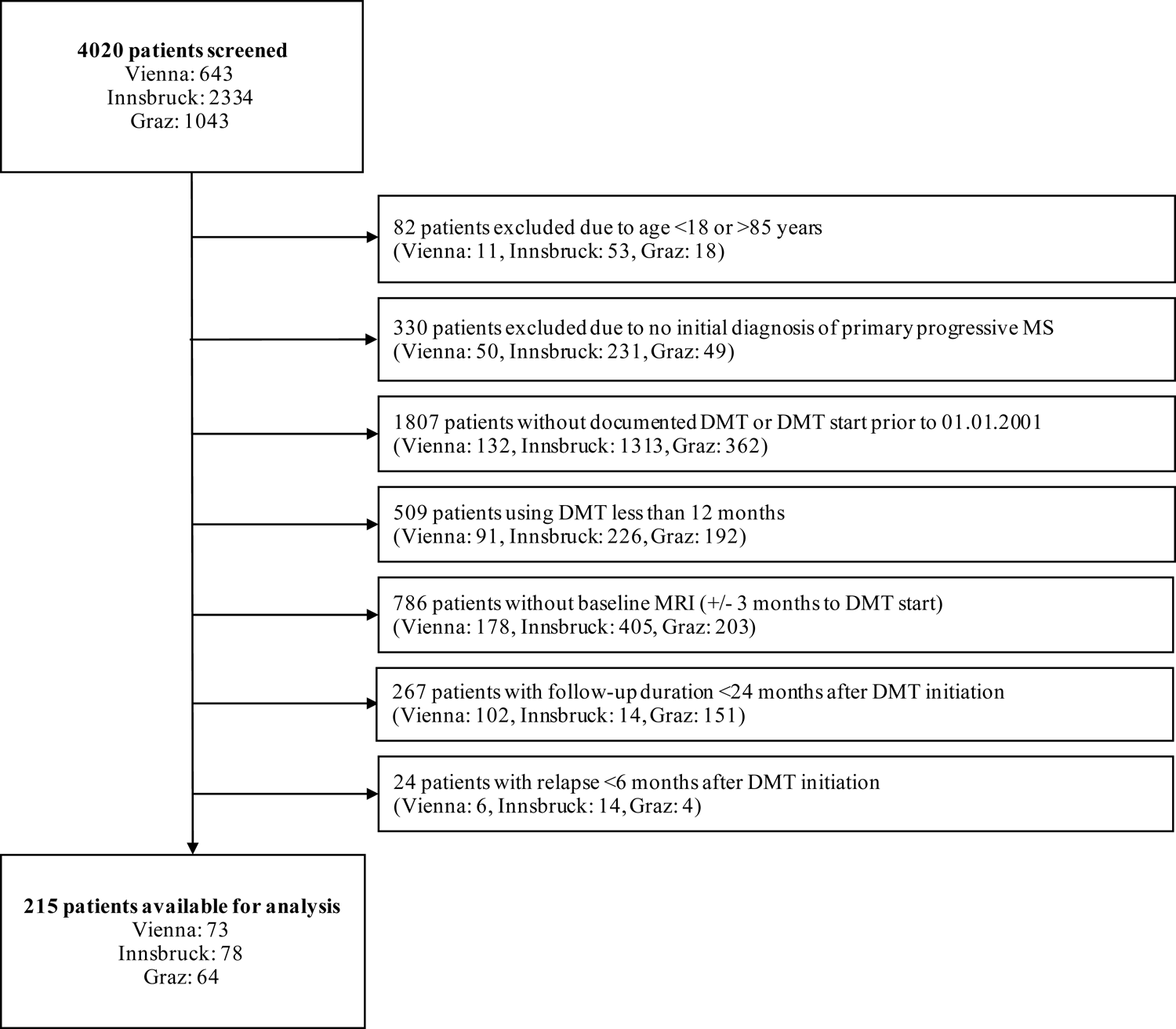
CONSORT Flow chart. *Abbreviations:* DMT = disease-modifying treatment, MS = multiple sclerosis

**Table 1:** Demographic, clinical and imaging characteristics.

|  | **Total cohort** | **Relapse during study period** | **No relapse during study period** | **p-value** |
| --- | --- | --- | --- | --- |
| **Number of patients** | 215 | 81 | 134 |  |
| **Age (years)** | 41<br>(32-53) | 38<br>(31-49) | 42<br>(33-54) | 0.128<sup>§</sup> |
| **Sex (female)** | 142<br>(66) | 61<br>(75) | 81<br>(60) | 0.037<sup>#</sup> |
| **Disease duration at DMT start (months)** | 2<br>(1-7) | 2<br>(1-6) | 3<br>(1-7) | 0.835<sup>§</sup> |
| **EDSS at DMT start** | 1<br>(0-2) | 1<br>(0-2) | 1<br>(0-2) | 0.665<sup>§</sup> |
| **≥9 T2 MRI lesions at DMT start** | 124<br>(58) | 53<br>(65) | 71<br>(53) | 0.099<sup>#</sup> |
| **≥1 CEL MRI lesions at DMT start** | 71<br>(33) | 28<br>(35) | 43<br>(32) | 0.822<sup>#</sup> |
| **OCB positivity<sup>1</sup>** | 185<br>(96) | 69<br>(97) | 116<br>(95) | 0.712<sup>*</sup> |
| **Number of relapses 12 months prior DMT start** | 1<br>(1-1) | 1<br>(1-1) | 1<br>(1-1) | 0.849<sup>§</sup> |
| **Moderate-efficacy DMT** | 159<br>(74) | 66<br>(81) | 93<br>(69) | 0.072<sup>2</sup> |
| **High-efficacy DMT** | 56<br>(26) | 15<br>(19) | 41<br>(31) |  |

Data are given as median (25<sup>th</sup>-75<sup>th</sup> percentile) and n (%). Comparisons were done by <sup>#</sup> $\chi^2$ test, <sup>\*</sup>Fisher's exact test and <sup>§</sup>Mann-Whitney U test.
<sup>1</sup> Results of OCB available of 193 patients (71 in the relapse activity group, 122 in the non-relapse activity group)
<sup>2</sup> Frequency of moderate vs. high-efficacy DMT in patients with and without relapse activity were compared.
*Abbreviations:* CEL = contrast-enhancing lesion, DMT = disease modifying treatment, EDSS = Expanded Disability Status Scale, MRI = magnetic resonance imaging, OCB = oligoclonal bands.

Patients were observed until first relapse or up to end of follow-up with a median time of 42 (28-58) months of observation. Eighty-one (38%) patients had relapse after a median of 28 (13-45) months. Univariate comparisons between patients with and without relapses are given in Table 1. Forty-six (21%) patients showed disability accrual after a median 34 (24-53) months. Univariate comparisons between patients with and without disability accrual are given in e Table 3.

### Efficacy of DMT in preventing relapse activity depended on patients’ age

The risk of relapse depended on both patients’ age and DMT strategy (Table 2). Higher age was associated with decreased risk of relapse (hazard ratio [HR]: 0.95 per year, 95% confidence interval [CI]: 0.93-0.98, p<0.001). The use of H-DMT lowered the risk of relapse compared to M-DMT (HR 0.06, 95% CI: 0.01-0.45, p=0.007). However, the benefit of H-DMT compared to M-DMT was reduced by increasing age (HR: 1.06 per year, 95%-CI: 1.01-1.11, p=0.032) (Figure 2). Superiority of H-DMT over M-DMT was estimated to be lost at the age of approximately 50 years. The remaining covariates, i.e., sex, disease duration, EDSS score at baseline, the number of relapses in the prior year as well as the number of T2L had no statistically significant impact on the risk of relapse (Table 2).

**Figure 2.**
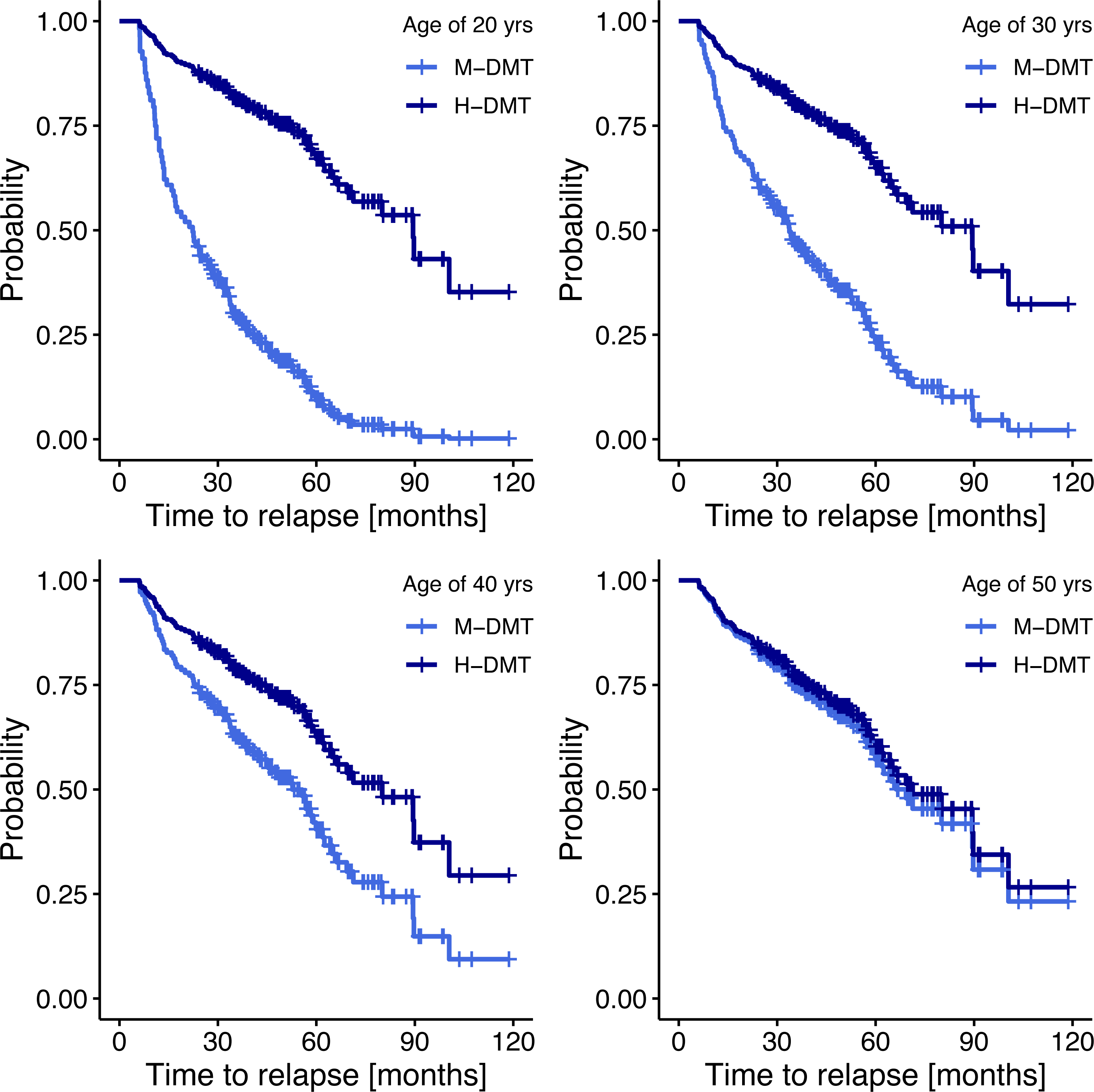
Probabilities of relapse depending on type of disease-modifying treatment at different patients’ age. The estimated probabilities for freedom of relapse depending on the type of DMT (M-DMT vs. H-DMT) for different patients’ ages (20, 30, 40, 50) are derived from the Cox regression model (Table 2). Adjusting co-variables were set as follows: sex to “female”, T2L to “≥9”, CEL to ≥1; and all other parameters to their median values. *Abbreviations*: M-DMT = moderate-efficacy disease-modifying treatment. H-DMT = moderate-efficacy disease-modifying treatment

**Table 2:** Cox regression predicting the risk of relapse dependent on disease-modifying treatment and age.

|  | Coefficient | Standard Error | P value | Hazard Ratio | 95%-CI |  |
| --- | --- | --- | --- | --- | --- | --- |
| **DMT** (ref: M-DMT) | -2.904 | 1.069 | **0.007** | **0.055** | 0.007 | 0.445 |
| **Age** (years) **at DMT start** | -0.048 | 0.012 | **<0.001** | **0.953** | 0.931 | 0.976 |
| **Sex** (ref: male) | 0.493 | 0.273 | 0.071 | 1.637 | 0.958 | 2.797 |
| **EDSS at DMT start** | 0.096 | 0.094 | 0.305 | 1.101 | 0.916 | 1.324 |
| **Disease duration** (months) **at DMT start** | 0.001 | 0.004 | 0.764 | 1.001 | 0.993 | 1.010 |
| **Number of relapses 12 months prior to DMT start** | 0.123 | 0.214 | 0.565 | 1.131 | 0.744 | 1.720 |
| **T2 MRI lesions at DMT start** (ref: <9) | 0.357 | 0.253 | 0.159 | 1.429 | 0.870 | 2.348 |
| **CEL MRI lesions at DMT start** (ref: 0) | 0.045 | 0.251 | 0.858 | 1.046 | 0.639 | 1.712 |
| **H-DMT : Age** | 0.056 | 0.026 | **0.031** | **1.058** | 1.005 | 1.113 |

Cox Snell pseudo-R<sup>2</sup>: 0.28
*Abbreviations:* CEL, contrast-enhancing lesions, CI = confidence interval, DMT = disease modifying treatment, EDSS = Expanded Disability Status Scale, H-DMT = high-efficacy disease modifying treatment, M-DMT = moderate-efficacy disease modifying treatment, MRI = magnetic resonance imaging, ref = reference categor

With regard to the risk of disability accrual, neither DMT strategy nor age nor the interaction term of DMT and age were found to be statistically significant predictors (eTable 4).

## Discussion

Our objective was to examine, in a DMT-naïve real-world cohort of RMS patients aged from 21 to 73 years, i) whether the advantage of initiating H-DMT over M-DMT in reducing relapse risk decreases with age, and ii) if so, at what age this superiority is lost.

Our findings indicate that older age is associated with both a reduced relapse risk and a diminished relative benefit of H-DMT over M-DMT in preventing relapses. Notably, in patients initiating their first DMT at approximately 50 years of age or older, the superior efficacy of H-DMT over M-DMT was no longer evident.

While the mechanisms underlying focal inflammation remain consistent across different age groups, the level of clinical and radiological disease activity – which forms the basis for DMT efficacy – declines with increasing age ^3–6^. The natural decrease in disease activity reduces the potential relative added value of DMT in patients at higher age (Figure 3). This is in line with age-stratified post-hoc analyses from randomized controlled trials, where the relative efficacy of both H-DMT and M-DMT is consistently strongest in the youngest age groups gradually decreasing with advancing age ^8,9,17^. A meta-analysis of multiple randomized controlled DMT trials found that the relative benefit of H-DMT over M-DMT regarding disability progression vanishes in patients over the age of 40-50 years, whereas another meta-analysis of largely the same trials using similar methodology showed that the reduction of relapses and MRI lesions did not significantly decrease with age ^7,10^. Clinical trial populations are typically limited to patients aged between 18 and 50 years and often have selection criteria that favour patients with high baseline disease activity. Consequently, these trials do not fully represent the diverse spectrum of patients in real-world practice ^10^. While it may be assumed that these trends continue in older patients, this has yet to be confirmed ^7–9^.

**Figure 3:**
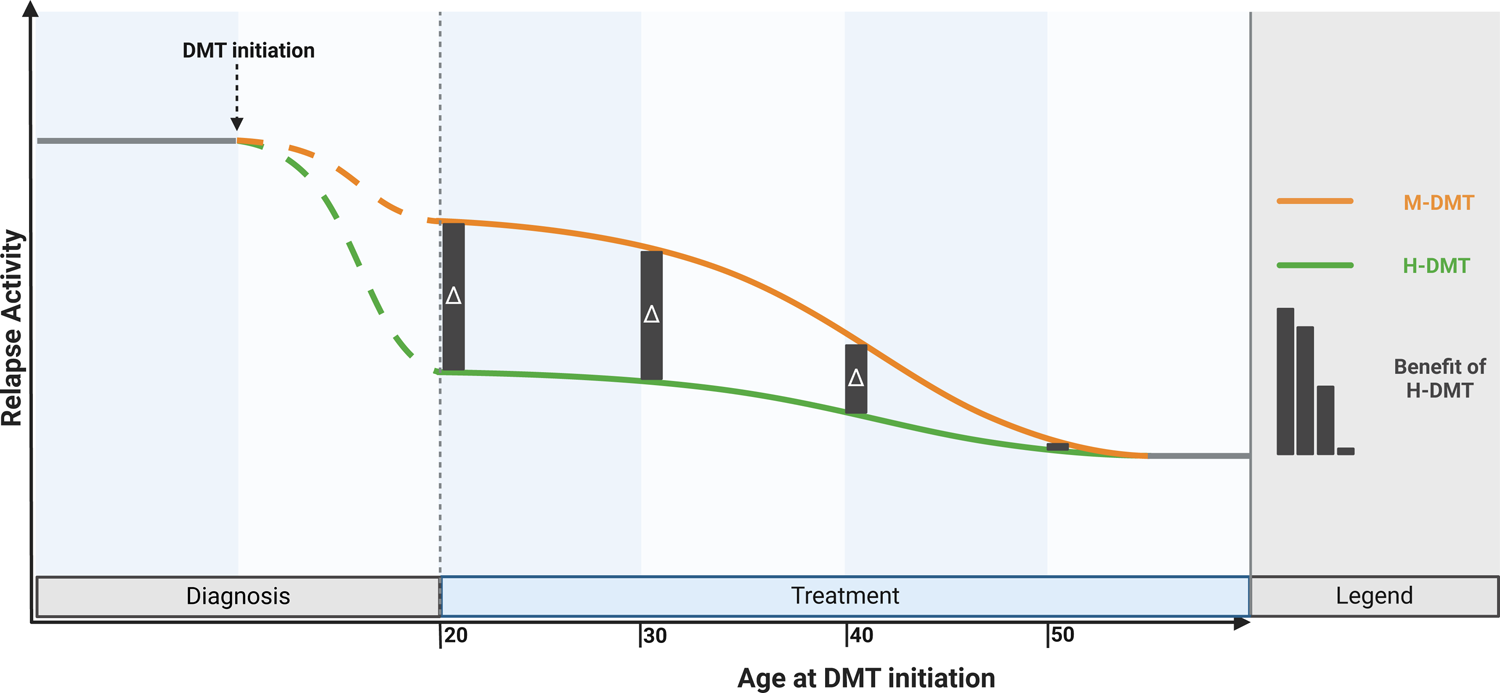
Conceptual impact of age on efficacy of first-line disease-modifying treatment in multiple sclerosis *Abbreviations*: DMT = disease-modifying treatment

On the contrary, the risk of adverse events from DMTs, particularly infectious complications, increases in older patients (associated with a growingly senescent immune system), and the higher prevalence of comorbidities such as cardiovascular and metabolic diseases complicates treatment ^18,19^.

Current treatment recommendations often do not emphasize the role of age as an independent predictor of treatment response and risk. Although approaches are increasingly moving towards the default use of H-DMT over M-DMT, older patients may have less need for highly potent therapies due to the natural decline in inflammatory disease activity with age. This, coupled with the heightening risk of treatment-related adverse events in older individuals, underscores the importance of developing age-specific treatment strategies ^2,20–22^.

In light of our findings, an age of approximately 50 years may serve as a reasonable threshold beyond which first-line use of H-DMTs should not be the default approach. Instead, for patients over this age, treatment decision should be made on an individual basis, following a rigorous assessment of the benefit-risk ratio.

However, it needs to be emphasized that our study does not imply that patients older than 50 years should not receive H-DMT. Our model is based on the average outcome within the study cohort, whereby an individual patient’s position within this distribution cannot be inferred from group data. Indeed, a patient older than 50 years suffering relapses and showing high T2L load with numerous CELs, i.e., above-average disease activity, is likely to then derive above-average benefit from DMT and may therefore well qualify for first-line use of H-DMT. However, such patients are infrequent in clinical routine. Conversely, existing prognostic tools lack the precision to accurately determine the degree of DMT efficacy required to suppress disease activity in individual patients. Monitoring methods can indicate only the presence or absence of disease activity on a given DMT but are unable to ascertain whether a DMT with lower efficacy would sufficiently control disease activity for a specific patient ^23^. Applying H-DMT universally would result in substantial overtreatment and expose patients to unnecessary treatment-related risks ^24^. This is especially concerning in older patients, for whom DMT associated risks are significantly greater ^18,19^. If every patient over 50 years were prescribed first-line H-DMT, our study suggests that half of these patients would be exposed to a cumulative risk for adverse events with little or no therapeutic benefit. Therefore, the tremendous interindividual heterogeneity of MS beyond the influence of age requires tailoring treatment decisions individually in order to achieve an optimal balance between suppression of disease activity and avoiding overtreatment with increased risk of adverse events ^25,26^. In our view, this would mean that the majority of DMT-naïve RMS patients older than age 50 should start with M-DMT, with close monitoring through regular clinical assessments and imaging. Upon the emergence of clinical or radiological evidence of ongoing disease activity, prompt switching to H-DMT is then required ^27^.

While it is well established that clinical and radiological disease activity decrease both with older age and longer disease duration, it remains unclear whether this decline is primarily driven by aging itself or whether disease duration independently contributes to this patho-physiological shift ^5^. Given the inherent correlation between age and disease duration, distinguishing their individual effects is challenging. However, in the present study, disease duration is highly unlikely to have played a major role, as the median duration at DMT initiation was only two months (range: 1–7 months).

With regard to disability accrual, we did not observe a statistically significant effect of DMT due to age. While our study was powered for the primary endpoint risk of relapse, the numbers of patients and observed events were probably too low to detect a statistically significant effect for the secondary endpoint disability accrual. However, the coefficients for DMT and the interaction term DMT:age allow a similar descriptive interpretation, i.e., one might conclude that the impact of DMT on disability accrual also diminishes with increasing age, even though, the overall effect sizes are lower. This is in line with most clinical trials, where treatment effects are typically lower on disability accrual than on relapse risk ^8,9,17^.

The strengths of this study lie in the high-quality data within the AMSD stemming from harmonized specialized MS centers in Austria with close-meshed follow-up providing a well-characterized cohort ^11,28–30^. Nevertheless, our study has several limitations. First, the sample size of the study is moderate, likely leading to underpowering regarding the secondary endpoint, wherefore further studies including a higher number of patients are necessary to substantiate our findings. Second, the retrospective analyses of data collected in clinical routine create possible confounders. An indication and/or selection bias cannot be ruled out, as DMT choice was made independent of the study and without randomization. However, the study was purposefully designed to assess whether age affects DMT response, aligning this population with the real-world clinical setting where decisions on initiating M-DMT or H-DMT occur. It’s important to note that our findings are specifically applicable to DMT-naïve patients and not to decisions about switching DMT in case of ongoing disease activity. In such scenarios, the persistent disease activity itself would likely necessitate a switch to H-DMT regardless of age. In addition, we were unable to consider ethnicity in our analysis, as our study population comprises >95% Caucasians. Future research should address this limitation. Finally, while the endpoint of time to relapse is robust and widely adopted in RMS, it does not reflect the entire spectrum and consequences of MS associated pathology, which likely also influences long-term clinical outcome.

The field’s understanding of DMT efficacy has been largely limited to patients aged between 18 and 50 years. The present study extends that knowledge by providing evidence on comparative DMT efficacy in a cohort of patients up to approximately 70 years of age. Our findings indicate that the superiority of H-DMT over M-DMT is lost in patients above approximately 50 years of age. These results underscore the urgent need for future clinical trials on DMT to include an appropriate representation to allow for a more comprehensive evaluation of DMT efficacy and associated risk across specific age groups ^18,31^. Future research should aim to explore how age can be integrated into personalized predictions of treatment response and risk profiles, potentially leading to a more individualized treatment strategies in patients with MS across all age groups.

## Data Availability

Anonymized data supporting the findings of this study are available from the corresponding au-thor upon reasonable request by a qualified researcher and upon approval by the data-clearing committee of the Medical Universities of Vienna.

## Acknowledgement

We thank all the AMSD investigators, clinical research staff, and especially the patients for helping to collect these data. The named individuals were not compensated for their help.

## Group information Austrian Multiple Sclerosis Database

AMSD investigators in alphabetical order: Altmann, Patrick (Medical University of Vienna, Vienna, Austria); Auer, Michael (Medical University of Innsbruck, Innsbruck, Austria); Berek, Klaus (Medical University of Innsbruck, Innsbruck, Austria); Barket, Robert (Medical University of Innsbruck, Innsbruck, Austria); Berger, Thomas (Medical University of Vienna, Vienna, Austria); Bsteh, Gabriel (Medical University of Vienna, Vienna, Austria); Damulina, Anna (Medical University of Graz, Graz, Austria); Deisenhammer, Florian (Medical University of Innsbruck, Innsbruck, Austria); Enzinger, Christian (Medical University of Graz, Graz, Austria); Di Pauli, Franziska (Medical University of Innsbruck, Innsbruck, Austria); Hegen, Harald (Medical University of Innsbruck, Innsbruck, Austria); Heschl, Bettina (Medical University of Graz, Graz, Austria); Khalil, Michael (Medical University of Graz, Graz, Austria); Kornek, Barbara (Medical University of Vienna, Vienna, Austria); Leutmezer, Fritz (Medical University of Vienna, Vienna, Austria); Monschein, Tobias (Medical University of Vienna, Vienna, Austria); Pinter, Daniela (Medical University of Graz, Graz, Austria); Rommer, Paulus (Medical University of Vienna, Vienna, Austria); Ropele, Stefan (Medical University of Graz, Graz, Austria); Schmidauer, Martin (Medical University of Innsbruck, Innsbruck, Austria); Schmied, Christiane (Medical University of Vienna, Vienna, Austria); Wurth, Sebastian (Medical University of Graz, Graz, Austria); Zebenholzer, Karin (Medical University of Vienna, Vienna, Austria); Zinganell, Anne (Medical University of Innsbruck, Innsbruck, Austria); Zulehner, Gudrun (Medical University of Vienna, Vienna, Austria); Zrzavy, Tobias (Medical University of Vienna, Vienna, Austria).

## Appendix 1 author contributions

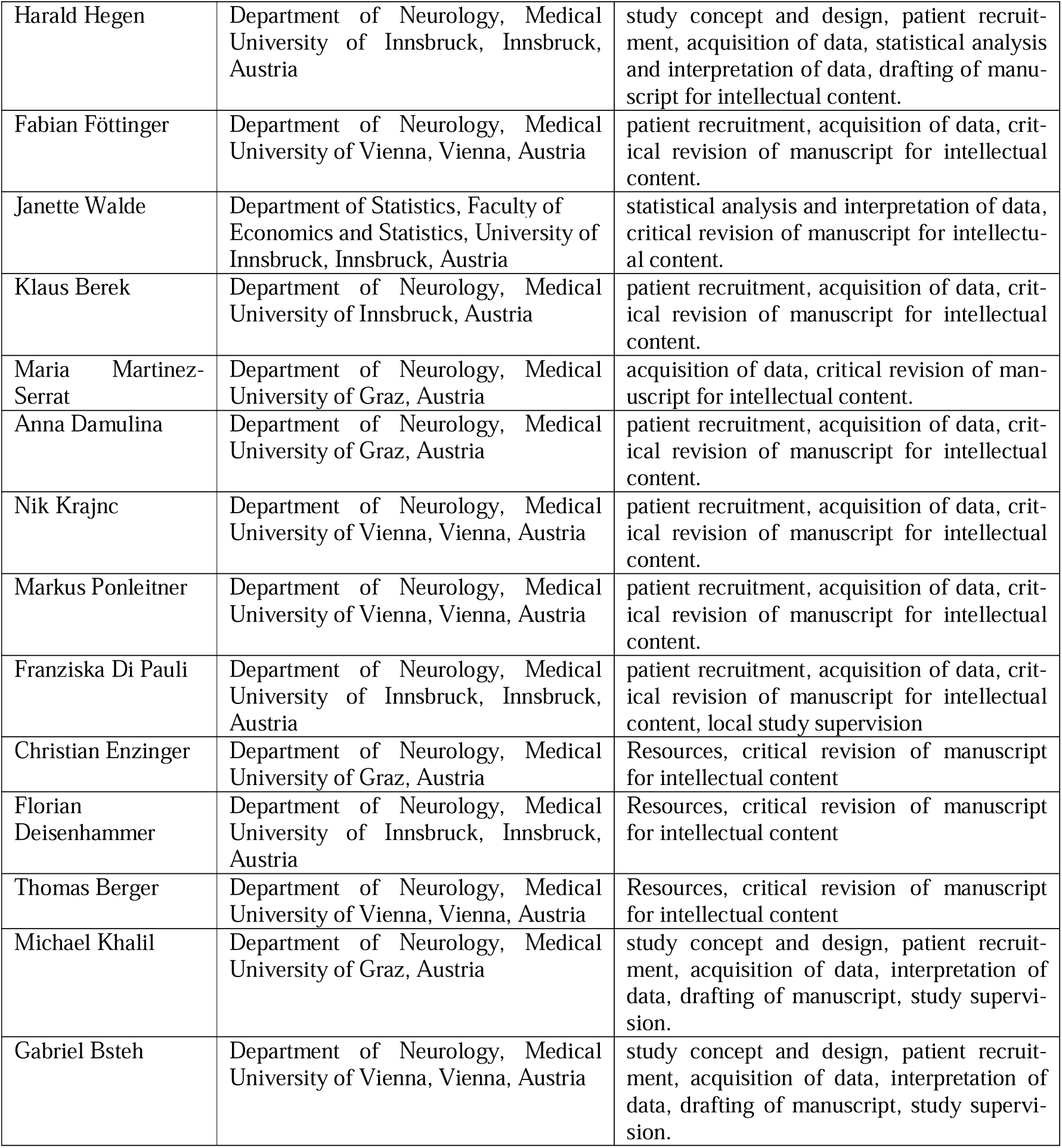

