## Supplemental Tables 1-4 for "The role of age in choosing high-efficacy treatment for multiple sclerosis – an Austrian MS Database study"

**Supplemental Table 1**: Different disease-modifying treatments applied in the study cohort

|  | **Total cohort** | **Relapse activity** | **No relapse activity** |
| --- | --- | --- | --- |
| **Moderate-efficacy DMT** | 159  (74) | 66  (81) | 93  (69) |
| Interferon-beta | 53  (25) | 23  (28) | 30  (22) |
| Glatiramer acetate | 31  (14) | 14  (17) | 17  (13) |
| Dimethyl fumarate | 68  (32) | 25  (31) | 43  (32) |
| Teriflunomide | 7  (3) | 4  (5) | 3  (2) |
| **High-efficacy DMT** | 56  (26) | 15  (19) | 41  (31) |
| Fingolimod | 9  (4) | 4  (5) | 5  (4) |
| Ozanimod | 4  (2) | 0  (0) | 4  (3) |
| Cladribine | 9  (4) | 3  (4) | 6  (4) |
| Natalizumab | 6  (3) | 1  (1) | 5  (4) |
| Rituximab | 20  (9) | 5  (6) | 15  (11) |
| Ocrelizumab | 5  (2) | 1  (1) | 4  (3) |
| Ofatumumab | 1  (0.5) | 0  (0) | 1  (1) |
| Alemtuzumab | 2  (2) | 1  (1) | 1  (1) |

Legend:

Data are given as n (%).

*Abbreviations*: DMT = disease-modifying treatment

**Supplemental Table 2:** Demographic, clinical and imaging characteristics per center

|  | **Graz** | **Innsbruck** | **Vienna** |
| --- | --- | --- | --- |
| **Number of patients** | 64 | 78 | 73 |
| **Age (years)** | 33  (27-41) | 37  (32-44) | 55  (49-60) |
| **Sex (female)** | 52  (80) | 47  (60) | 43  (60) |
| **Disease duration (months)** | 3  (1-7) | 2  (1-5) | 2  (1-8) |
| **EDSS at DMT start** | 1  (0-2) | 1  (0-1.5) | 2  (1-2.5) |
| **≥9 T2 lesions at DMT start** | 30  (46) | 42  (54) | 52  (72) |
| **≥1 CEL** | 24  (38) | 30  (38) | 17  (23) |
| **OCB positivity^1^** | 57  (92) | 70  (99) | 58  (97) |
| **Number of relapses 12 months prior DMT start** | 1  (1-1) | 1  (1-1) | 1  (1-1) |

Legend:

Data are given as median (25^th^-75^th^ percentile) and n (%).

^1^ Results of OCB available of 62 patients in Graz, of 71 in Innsbruck and 60 patients in Vienna.

*Abbreviations*: CEL = contrast-enhancing lesion, DMT = disease modifying treatment, EDSS = Expanded Disability Status Scale, OCB = oligoclonal bands.

**Supplemental Table 3:** Demographic, clinical and imaging characteristics

|  | **Total cohort** | **Disability accrual** | **No disability accrual** | **p-value** |
| --- | --- | --- | --- | --- |
| **Number of patients** | 215 | 46 | 169 |  |
| **Age (years)** | 41  (32-53) | 46  (37-55) | 38  (32-52) | 0.002^§^ |
| **Sex (female)** | 142  (66) | 32  (70) | 110  (65) | 0.694^#^ |
| **Disease duration (months)** | 2  (1-7) | 3  (1-9) | 2  (1-6) | 0.526^§^ |
| **EDSS at DMT start** | 1  (0-2) | 1  (0-2) | 1  (0-2) | 0.711^§^ |
| **≥9 T2 lesions at DMT start** | 124  (58) | 29  (63) | 95  (56) | 0.507^#^ |
| **≥1 CEL** | 71  (33) | 15  (33) | 56  (33) | 1.000^*^ |
| **OCB positivity^1^** | 185  (96) | 39  (93) | 146  (97) | 0.507^#^ |
| **Number of relapses 12 months prior DMT start** | 1  (1-1) | 1  (1-1) | 1  (1-1) | 0.510^§^ |
| **Moderate-efficacy DMT** | 159  (74) | 35  (76) | 124  (73) | 0.855^2^ |
| **High-efficacy DMT** | 56  (26) | 11  (24) | 45  (27) |  |

Legend:

Data are given as median (25^th^-75^th^ percentile) and n (%). Comparisons were done by ^#^χ^2^ test, ^*^Fisher’s exact test and ^§^Mann-Whitney U test.

^1^ Results of OCB available of 193 patients (39 in the disability accrual group, 146 in the non-disability accrual group)

^2^ Frequency of moderate vs. high-efficacy DMT in patients with and without disability accrual were compared.

*Abbreviations*: CEL = contrast-enhancing lesion, DMT = disease modifying treatment, EDSS = Expanded Disability Status Scale, OCB = oligoclonal bands.

**Supplemental** **Table 4**: Cox regression predicting the risk of disability progression

|  | **Coefficient** | **Standard Error** | **P value** | **Hazard Ratio** | **95%-CI** | |
| --- | --- | --- | --- | --- | --- | --- |
| **DMT** (ref: M-DMT)_ | -1.947 | 1.398 | 0.164 | 0.143 | 0.009 | 2.210 |
| **Age** (years) | 0.004 | 0.017 | 0.808 | 1.004 | 0.971 | 1.038 |
| **Sex** (ref: male) | 0.241 | 0.334 | 0.471 | 1.272 | 0.661 | 2.447 |
| **EDSS at DMT start** | 0.020 | 0.127 | 0.873 | 1.021 | 0.795 | 1.310 |
| **Disease duration** (months) | 0.002 | 0.005 | 0.680 | 1.002 | 0.993 | 1.011 |
| **Number of relapses 12 months prior**  **to DMT start** | 0.423 | 0.297 | 0.155 | 1.527 | 0.853 | 2.735 |
| **T2 lesions at DMT start** (ref: <9) | -0.115 | 0.338 | 0.734 | 0.892 | 0.460 | 1.729 |
| **CEL at DMT start** (ref: 0) | -0.170 | 0.342 | 0.619 | 0.844 | 0.431 | 1.649 |
| **H-DMT : Age** | 0.047 | 0.030 | 0.119 | 1.048 | 0.988 | 1.112 |

Cox Snell pseudo-R^2^: 0.16

*Abbreviations*: CEL, contrast-enhancing lesions, CI = confidence interval, DMT = disease modifying treatment, EDSS = Expanded Disability Status Scale, H-DMT = high-efficacy disease modifying treatment, M-DMT = moderate-efficacy disease modifying treatment, ref = reference category
